# Preconception weight change and pregnancy, birth, and child outcomes: Design and baseline characteristics of the MatTrack Cohort

**DOI:** 10.64898/2026.08.25.26361309

**Authors:** Meghan Mayhew, Kimberly K. Vesco, Deborah Rohm Young, Caryn Oshiro, Lasha S. Clarke, Ning Smith, Erin S. LeBlanc, Ashli A. Owen-Smith, Courtney E. McCraken, Michael C. Leo, Mi H. Lee, Botao Zhou, Carmen Wong, Alexander Hudgins, Natalie A. Rosenquist, Janne Boone-Heinonen

## Abstract

Current recommendations suggest that women achieve a healthy weight before becoming pregnant, but evidence supporting the benefits of preconception weight loss are limited and inconsistent, with some evidence of risks. The Maternal Preconception Weight Trajectory (MatTrack) study will evaluate the impact of preconception weight change on maternal, pregnancy, and child outcomes. In this paper, we present methods used to construct the cohort and evaluate baseline characteristics.

The MatTrack cohort was derived from electronic health record data from four Kaiser Permanente regions. Inclusion criteria addressed data quality, availability, and enrollment; maternal age (≥18 years); and date (pregnancy onset date in 2006-2020, delivery ≤12/31/2020). Starting body mass index (BMI) was calculated using weight closest to 24 months prior to pregnancy onset date and adult height. Preconception weight change rates from 24 months prior to and through pregnancy onset date were estimated using linear mixed effects models. Descriptive analyses characterized the baseline characteristics of the cohort.

The cohort includes 297,592 pregnancies with the full spectrum of starting BMI: underweight (2.3%), normal weight (41.8%), overweight (28.4%); and obesity class I (15.3%), II (7.4%), and III (5.0%). The cohort is demographically diverse, with 8.3% covered by Medicaid; and 44.5%, 9.2%, and 10.9% Hispanic, non-Hispanic Black, or non-Hispanic Asian, respectively.

Preconception weight change rates (kg/year) span weight loss to gain, with the greatest loss in those with obesity class III [median (10^th^, 90^th^ percentile): −0.8 (−9.5, 4.9)] and the greatest gain in those with underweight [median (10^th^, 90^th^ percentile): 1.1 (−0.7, 3.6)].

Longitudinal data from this cohort of nearly 300,000 linked maternal-child dyads will enable examination of associations between preconception weight loss and pregnancy, maternal, and child health outcomes. Findings will strengthen the evidence base for preconception weight management guidelines.

## Introduction

Maternal weight status prior to conception is a well-established risk factor for adverse pregnancy, birth, and child outcomes. Specifically, greater body mass index (BMI) at pregnancy onset is associated with higher risk of adverse pregnancy outcomes, including gestational diabetes (GDM), hypertensive disorders of pregnancy, and large-for-gestational-age delivery; [1] and child outcomes, particularly obesity. [2] Women who have underweight at conception (BMI <18.5 kg/m^2^) also have greater risk for adverse pregnancy, including preterm delivery, small-for-gestational-age (SGA), and low birth weight. [3, 4]

Thus, clinical practice guidelines recommend that women achieve a normal BMI (18.5 - 24.9 kg/m^2^) before pregnancy, corresponding to preconception weight loss for those with BMI ≥25 and preconception weight gain for women with BMI <18.5. [5, 6] However, evidence suggesting benefits of preconception weight change is limited and inconsistent [7, 8] and the timing, amount, and rate of preconception weight change that optimizes maternal and fetal health is largely unexplored. Further, emerging evidence suggests that preconception weight loss may be associated with greater gestational weight gain (GWG), [9, 10] raising concerns about weight rebound during pregnancy and potential unintended consequences of weight change prior to pregnancy.

The objective of the *Maternal Preconception Weight Trajectory (MatTrack)* study is to examine the effect of maternal preconception weight change on pregnancy, maternal, and child health outcomes in a large U.S. patient population. We developed a large cohort of pregnancies using 15 years of electronic health record (EHR) data from four U.S. health care systems and examined weight changes in the 24 months prior to pregnancy among patients spanning the full BMI spectrum. In this paper, we describe the methods used to assemble the MatTrack cohort, how key variables were defined, and baseline demographic and clinical characteristics of the cohort.

## Materials and Methods

### Study setting

The MatTrack study cohort includes patients with one or more pregnancies who received health care services at Kaiser Permanente Georgia, Hawaii, Northwest, or Southern California (KPGA, KPHI, KPNW, KPSC) in 2006-2020. Kaiser Permanente (KP) is a large, multi-regional integrated healthcare system with a comprehensive electronic health record (EHR) Virtual Data Warehouse (VDW), which provides standardized information from outpatient encounters, hospital admissions, laboratory results, prescriptions, and claims, providing a comprehensive source of data about preconception, pregnancy, delivery, postpartum, and child health. The KP VDW maintains a pregnancy outcome episode table that was created using an algorithm developed by the KP Center for Effectiveness and Safety Research, a national research collaborative comprising KP’s research centers. [11] The pregnancy episode table contains one record per pregnancy for which an outcome exists and key characteristics of each pregnancy including: outcome type (e.g., livebirth, stillbirth, spontaneous abortion, ectopic pregnancy); pregnancy outcome and onset dates, estimated delivery date, last menstrual period, gestational age; delivery type (vaginal, assisted vaginal, and cesarean); maternal age; gravidity, parity; and data quality variables identifying imputed values, weak data, conflicting evidence or overlapping episodes. [12] Preconception, postpartum, child, and additional pregnancy data for the MatTrack cohort were obtained from diagnosis and procedure codes, laboratory, and other data from the KP VDW as well as State vital statistics records.

This research was reviewed and approved by the KPNW Institutional Review Board (IRB) on October 4, 2021. The KPNW IRB agreed to accept review authority and continuing oversight for this study from KP Georgia, KP Southern California and KP Hawaii under an Interregional Reliance Agreement.

### Cohort creation

To develop the MatTrack cohort (Fig 1), we identified 674,967 pregnancies from the KP Center for Effectiveness and Safety Research pregnancy outcome tables of the VDW for the four KP regions with pregnancy onset dates between 1/1/2006 and 12/31/2020, among patients 18 years and older at pregnancy onset date. These retrospective data were first accessed for the study purpose on February 22, 2022 and extracted as a limited dataset. We required robust pregnancy dating and follow-up (i.e., documented pregnancy onset and outcome data, known pregnancy outcome), and evidence of KP health plan enrollment within the preconception period (25 months prior to pregnancy onset date). We required at least one valid weight measured within 25 months prior to pregnancy onset date and at least one valid height measured from 16 years of age through 18 months (540 days) after the last pregnancy onset date (7,517 pregnancies excluded, 1.1%). To model preconception weight trajectory, we also required at least one valid weight measure within each of the following two periods: 0 to 365 days prior to pregnancy onset date and 366 to 730 days prior to pregnancy onset date (267,200 (40.0%) excluded). Finally, we excluded pregnancies that resulted in early (<20 weeks gestation) pregnancy loss (93,287 excluded, 23.3%) or that were not singleton, live births (9,371 excluded, 3.1%). Procedures for identifying valid measures are described below in Study Variables. The final MatTrack Cohort includes 297,529 pregnancies among 240,057 women.

**Fig 1.**
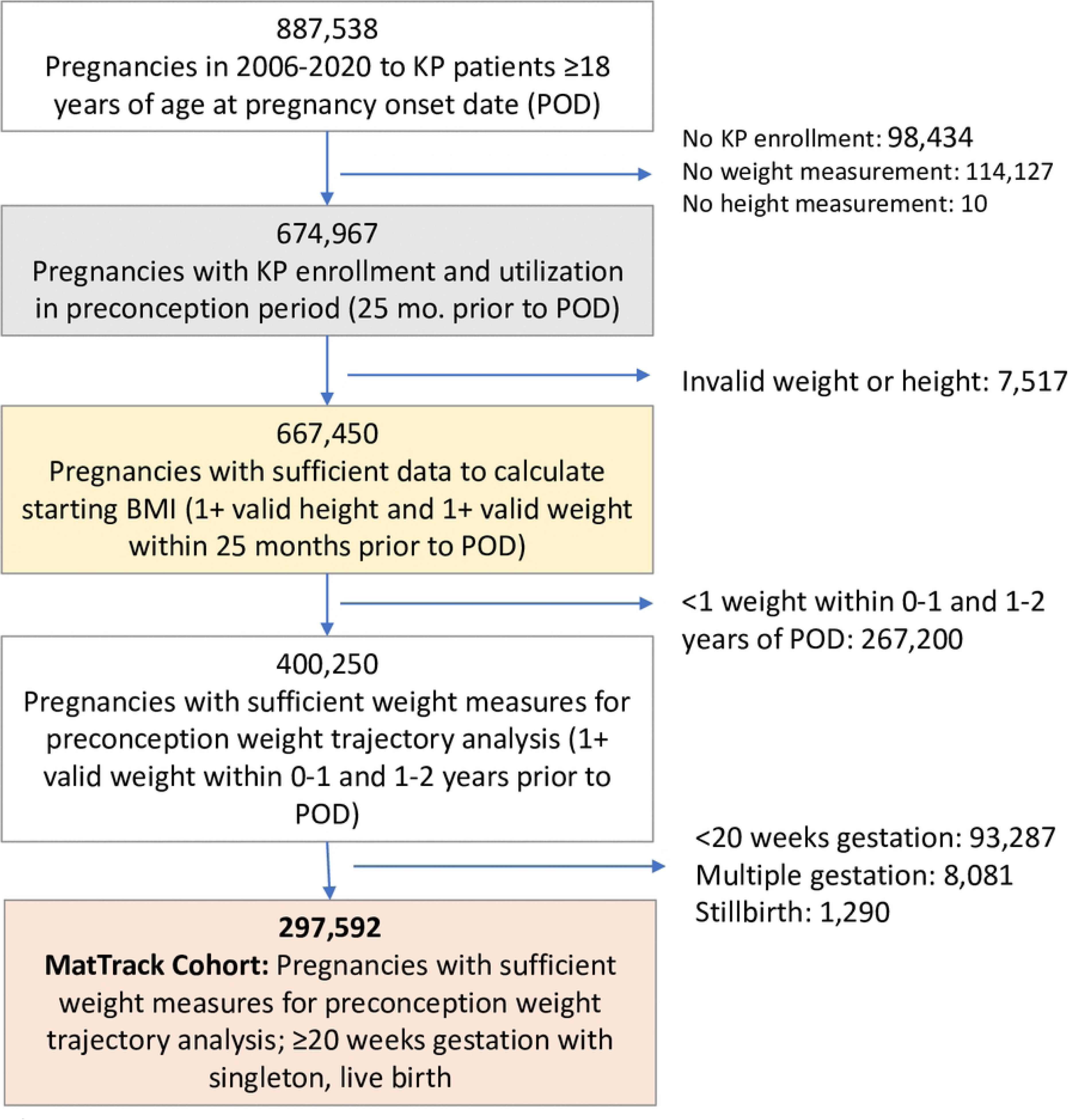
**Flow diagram describing identification of pregnancies for inclusion in MatTrack cohort**

### Linked data sources

#### Child clinical data

The KP VDW employs a standardized approach across KP regions to link KP maternal deliveries to corresponding infant KP EHR data using a validated deterministic algorithm. [13] Among the 297,529 pregnancies in the MatTrack Cohort, 97.1% were linked to the infants’ KP clinical records.

#### Birth certificate data

Each KP region obtained approval to utilize data elements from state-issued birth certificates for each state in which the participating KP regions reside: Hawaii for KPHI, Oregon and Washington for KPNW, California for KPSC, and Georgia for KPGA. Each KP site linked their *MatTrack Cohort pregnancies* to state birth records data. KPNW and KPGA used probabilistic linkage, matching identifier fields common to both records (name, DOB, etc.) using the SPEDIS function in SAS. KPHI and KPSC used deterministic linkage methods already established by each region. Among *MatTrack Cohort* pregnancies, 89.7% (N=267,007) were linked to state birth records data.

#### Identifying preconception maternal height and weights

Maternal preconception weight measures 36 months prior to pregnancy onset date and height measures from the 16^th^ birthday through 540 days (18 months) after the last observed pregnancy end date were extracted from clinical encounters. Weight and height measures were then screened [14] for implausible values using the adult module of the R growthcleanr package (v.2.2.0). [15, 16] growthcleanr identifies implausible weight and height measurements based on serial measurements within each individual patient and has been validated and tested in data from multiple health care systems. [15, 16] Minor customizations to growthcleanr for our study population are described in **S1 File**. Out of the 2,815,824 pre-pregnancy weight measures recorded within 36 months before pregnancy onset date, 3.4% were flagged and excluded. Similarly, 7.3% of the 9,679,454 maternal height measures were excluded. Among the remaining (valid/plausible) height and weight measures, we constructed the following maternal preconception variables.

#### Maternal height

Maternal height was defined as the median recorded height for each patient, among plausible values. Heights measured at 16 years or older aligned with the preconception period for the youngest cohort members (18 years at the start of pregnancy). We allowed heights starting at age 16 as most females will have reached their adult height by that time.[17, 18]

#### Starting BMI

We calculated starting BMI within the preconception period (0-24 months prior to pregnancy onset date) using height and the first observed, valid weight, among weights measured from >12 to 24 months (366 to 730 days) prior to pregnancy onset date. In the primary analysis, starting BMI was analyzed as a categorical stratification variable: Underweight (<18.5), Normal weight (18.5 to <25), Overweight (25 to <30), Obesity Class I (30 to <35), Obesity Class II (35 to <40), and Obesity Class III (≥40).

#### Preconception weight change (kg/year; exposure)

Preconception weight change was characterized by modeling all valid weight measures in the 24 months prior to pregnancy onset date using the longitudinal linear mixed effects model described in the Statistical Analysis section.

#### Covariates

Covariates were ascertained from two data sources: the EHR, which was the primary source for most variables, and state birth certificate data. Maternal age was calculated for the start of the pregnancy using mother’s date of birth and pregnancy onset date from the EHR. Tobacco use was ascertained from social history questionnaire data recorded in the EHR, defined as any instance of tobacco use recorded during the preconception period (0-24 months prior to pregnancy onset date) and during pregnancy (pregnancy onset to pregnancy outcome date). Insurance type was defined as the insurance type recorded in the earliest insurance enrollment period in the 25 months (760 days) prior to pregnancy onset date recorded in the VDW, classified as Public, Private or Other. Public insurance type was defined based on having any of the following coverage during the period: Medicaid or Medicare; otherwise, Private was defined based on having any of the following coverage: commercial or private pay; and Other was defined as any other insurance type, such as state-subsidized insurance. Parity was defined using state birth certificates (primary source; 89.3%) and EHR data (secondary source if no linked birth certificate; 10.3%) with 0.3% missing, dichotomized as nulliparous versus parous.

Race and ethnicity were defined using state birth certificate data as the primary source (89.1%) and EHR data as a secondary source (10.6%), with 0.3% missing from both sources. This approach was used because race and ethnicity are self-reported on state birth certificates, while the source of race and ethnicity information in the EHR varies by KP region and is not necessarily derived from individual patient surveys with detailed categorical options. We mapped race and ethnicity categories, which varied across state and over time, into standard categories, then created a combined race/ethnicity variable:^25^ Hispanic, non-Hispanic (NH) American Indian/Alaska Native, NH Asian, NH Black, NH Native Hawaiian/Other Pacific Islander, NH Other, NH More than one race, NH White, Unknown/not reported. People who self-identified as Hispanic were classified as such and then all other individuals were grouped using all race information provided. Individuals were classified as multiracial if they did not identify as Hispanic and identified with at least two races.

Neighborhood deprivation index (NDI) [19] for years 2012-2020 was available in the VDW for each recorded KP enrollee address. We extracted NDI for the Census Tract and calendar year of the participant’s address record of longest duration in the 24 months prior to POD [POD – 730 days]. For pregnancies with a POD prior to 2012 (2006-2011), the 2012 value was used. NDI was designated as missing if no address was recorded in the 24 months prior to POD or the address was not within a KP regional service area.

### Statistical analysis

In descriptive analysis, we examined sociodemographic and clinical characteristics across maternal starting BMI categories. Here we report frequencies and percentages for categorical variables and means and standard deviations (SD) for continuous variables.

Preconception weight change at the pregnancy level was estimated using the 3-level linear mixed effects model[20] where the outcome was all valid weight measures in the 24 months prior to POD. This model took into account two levels of clustering: repeated longitudinal weight measurements over time (Level 1) within pregnancy (Level 2), which were further nested within each individual patient (Level 3), i.e., ≥1 pregnancy per patient. The model included the following fixed effect terms: time in years as a continuous variable, starting BMI categories (categorical variable), and the interaction of starting BMI categories by time to allow different weight trajectories across starting BMI categories. No other covariates were adjusted in the model because the goal of this model was to characterize the preconception weight trajectory, not to identify association of preconception weight with other characteristics. In terms of random effects, a random intercept was specified at the patient level to accommodate correlated pregnancies within patients; and a random intercept and a random slope for time with an unstructured covariance matrix were included at the pregnancy level to allow individual pregnancy baseline and trajectory variability over time. All model parameters were estimated via restricted maximum likelihood (REML) using SAS PROC MIXED. The estimated fixed and random time slopes were then combined as the estimated absolute preconception weight change rate (kg/year) for each pregnancy. The distribution of the absolute preconception weight change rate was examined, and extreme values defined as ≥3 SD above or below the mean were identified and excluded[21] (n=4,331, 1.5% excluded). Additionally, annual weight change as a percentage of starting weight (used to calculate starting BMI, above) was calculated as *100*[preconception weight change rate (kg/year)]/[starting weight]*, hereafter referred to as “percent weight change rate.” Percent weight change rate was considered both as a continuous measure and also categorized as >5% loss, within 5%, or >5% gain, as changes in weight of 5% or greater are considered to be clinically relevant and weight maintenance is often defined as weight fluctuations of ≤5% of starting weight. [22]

We then conducted descriptive analysis of absolute and percent weight change rates, including calculation of mean, SD, and selected percentiles within starting BMI category and display of the full distribution with violin plots.

## Results

Of the 674,967 pregnancies with evidence of Kaiser Permanente Health Plan enrollment during the study period, 400,250 had the required number of weights to calculate a 2-year preconception weight trajectory (**Figure 1**). Of those, there were 297,592 that resulted in singleton, livebirth at ≥ 20 weeks gestation. We compared the characteristics of pregnancies in the final analytic cohort to the greater population of pregnancies identified prior to applying exclusion criteria and found the cohorts were similar with respect to starting BMI, smoking, and sociodemographic characteristics (**S1 Table**).

**Table 1** describes the characteristics of pregnancies included in the MatTrack analytic cohort. Overall, the MatTrack cohort includes a representative population of patients across the reproductive age and BMI spectrum. Patients had a mean age of 29.8 years, 60.8% were multiparous, most did not smoke prior to (92.1%) or during (96.0%) pregnancy, and the cohort was predominantly (91.7%) commercially insured. The largest racial and ethnic groups were Hispanic (44.5%), non-Hispanic white (29.8%), non-Hispanic Asian (10.9%), and non-Hispanic Black (9.2%).

**Table 1.** Baseline characteristics of the MatTrack cohort, by preconception starting BMI category^a^.

|  | Under-<br>weight<br>N=6,691 | Normal<br>N=124,364 | Overweight<br>N=84,368 | Obesity<br>Class I<br>N=45,459 | Obesity<br>Class II<br>N=21,952 | Obesity<br>Class III<br>N=14,758 | TOTAL<br>N=297,592 |
| --- | --- | --- | --- | --- | --- | --- | --- |
| **Age at pregnancy onset**<br>**[mean (SD)]** | 27.6 (5.9) | 29.5 (5.7) | 30 (5.5) | 30.2 (5.5) | 30.3 (5.4) | 30.5 (5.2) | 29.8 (5.6) |
| **Parity (%)** |  |  |  |  |  |  |  |
| Nulliparous | 57.2 | 46.0 | 34.4 | 31.3 | 31.3 | 34.7 | 39.1 |
| Multiparous | 42.4 | 53.7 | 65.3 | 68.4 | 68.4 | 65.0 | 60.6 |
| Missing | 0.4 | 0.4 | 0.3 | 0.3 | 0.3 | 0.4 | 0.3 |
| **Smoking prior to**<br>**pregnancy (%)** |  |  |  |  |  |  |  |
| No | 90.0 | 92.8 | 92.6 | 91.3 | 90.1 | 88.9 | 92.1 |
| Yes | 9.6 | 6.8 | 7.2 | 8.4 | 9.6 | 10.8 | 7.6 |
| Missing | 0.4 | 0.4 | 0.3 | 0.3 | 0.3 | 0.3 | 0.3 |
| **Smoking during**<br>**pregnancy (%)** |  |  |  |  |  |  |  |
| No | 94.7 | 96.5 | 96.3 | 95.6 | 94.8 | 94.1 | 96.0 |
| Yes | 5.0 | 3.2 | 3.5 | 4.2 | 5.0 | 5.7 | 3.7 |
| Missing | 0.3 | 0.3 | 0.2 | 0.2 | 0.2 | 0.2 | 0.3 |
| **Insurance type (%)** |  |  |  |  |  |  |  |
| Commercial | 90.5 | 93.6 | 92.1 | 89.8 | 87.8 | 85.8 | 91.7 |
| Public | 9.5 | 6.4 | 7.9 | 10.2 | 12.2 | 14.2 | 8.3 |
| **Maternal race/ethnicity**<br>**(%)** |  |  |  |  |  |  |  |
| Hispanic | 29.9 | 36.9 | 48.5 | 53.1 | 53.9 | 50.8 | 44.5 |
| NH White | 31.8 | 34.5 | 28.1 | 24.4 | 24.0 | 23.9 | 29.8 |
| NH Asian | 22.7 | 15.9 | 8.9 | 5.5 | 3.1 | 1.7 | 10.9 |
| NH NH/PI | 1.2 | 1.0 | 1.1 | 1.3 | 1.8 | 2.0 | 1.2 |
| NH Black | 9.4 | 7.1 | 9.1 | 11.0 | 12.5 | 16.6 | 9.2 |
| NH Multiracial | 3.3 | 3.3 | 3.3 | 3.6 | 3.9 | 4.1 | 3.4 |
| NH Other | 0.2 | 0.1 | 0.1 | 0.1 | 0.1 | 0.2 | 0.1 |
| Unknown | 1.6 | 1.1 | 0.9 | 0.9 | 0.7 | 0.8 | 1.0 |
| **Neighborhood**<br>**Deprivation Index**<br>**[mean (SD)]** | 0.2 (0.9) | 0.1 (0.9) | 0.3 (1.0) | 0.5 (1.0) | 0.6 (1.0) | 0.6 (1.0) | 0.3 (1.0) |

<sup>a</sup>Starting BMI categorized as Underweight (<18.5), Normal weight (18.5 to <25), Overweight (25 to <30), Obesity Class I (30 to
<35), Obesity Class II (35 to <40), and Obesity Class III (≥40 kg/m<sup>2</sup>)
BMI, Body Mass Index; NH, Non-Hispanic

Patients with preconception Class I to III obesity were more likely to be multiparous (65.0-68.4%), smoke prior to (8.4-10.8%) or during (4.2-5.7%) pregnancy, and be publicly insured (10.2-14.2%) (**Table 1**). Hispanic (50.8-53.9%) and non-Hispanic Black (11.0-16.6%) patients had greater representation in the obesity groups. Patients with preconception underweight were slightly younger (28.2 years), were more likely to smoke prior to (9.6%) or during (5.0%) pregnancy and be publicly insured (9.5%). Non-Hispanic Asian patients had a greater representation in the underweight group (22.7%). Neighborhood deprivation was higher with greater starting BMI (0.1 for normal weight vs. 0.6 for Class II and III obesity).

Average rate of preconception weight change during the 2-year preconception period was 0.4 kg/year gain (SD 3.4, IQR: 1.3 loss, 2.3 gain). Median weight change was lower with increasing starting BMI, from 1.1 kg/year gain among those with underweight to 0.6 kg/year gain in normal weight and 0.8 kg/year loss in obesity class III (**Fig 2 and S2 Table**). Preconception weight change rate was more variable with greater starting BMI, with IQRs increasing from 0.2 to 2.2 in underweight to −1.3 to 2.3 in obesity class III.

**Fig 2.**
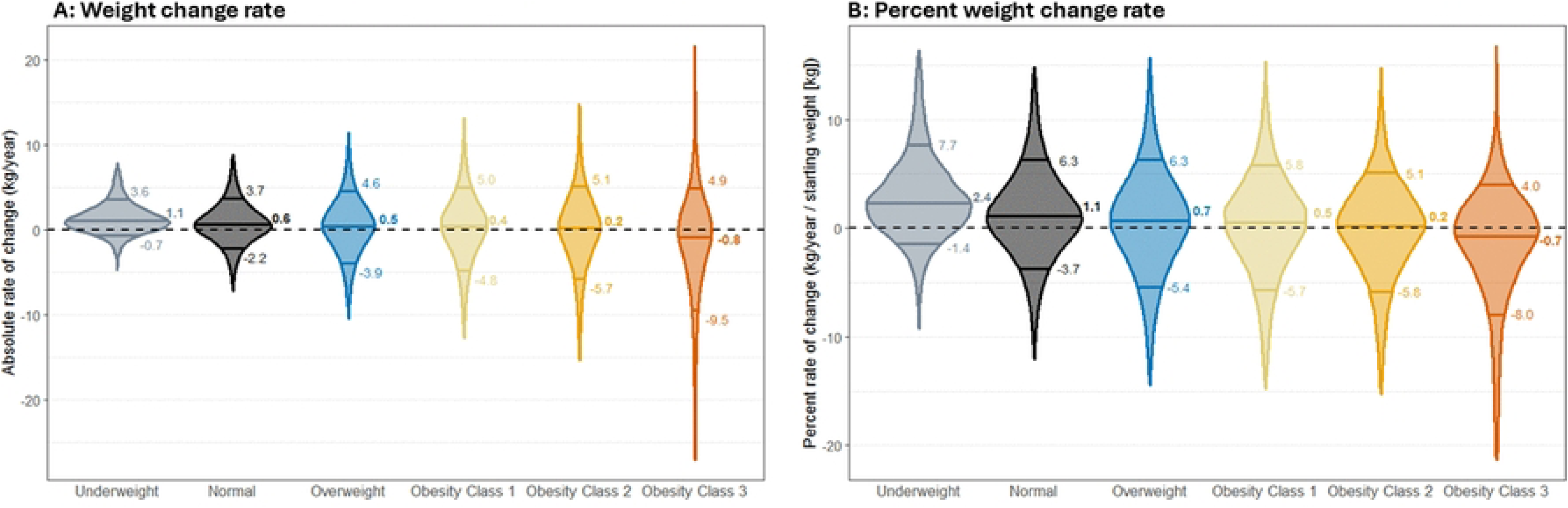
Distribution of the rate of preconception weight change (median, 10^th^, and 90^th^ percentiles*) as absolute weight change (A) and percent weight change (B) *The median is represented by the middle horizontal line; 10^th^ and 90^th^ percentiles are represented by the 1^st^ and 3^rd^ crosses; the length of each plot represents the observed minimum to maximum values.

When expressed as a percentage of starting weight, preconception weight change rate was on average 0.7% gain/year (SD 4.5, IQR: 1.8% loss, 3.3% gain) (**Fig 2 and S3 Table**). Median percent weight change rate was 2.4% gain per year in those with underweight, decreasing to 0.7% loss per year in those with obesity class III. Compared to absolute weight change rate, variability in percent weight change rate was more consistent across starting BMI categories (underweight: IQR (0.4%, 4.7%), SD (3.7%); obesity class III: IQR (−3.8%, 1.8%), SD (5.2%)). Overall, 9.4% lost >5% of their starting weight per year and 14.5% gained >5% of their starting weight per year, with the highest proportion with >5% loss in the obesity class III group (19.0%).

## Discussion

The MatTrack cohort includes nearly 300,000 pregnancies with detailed characterization of maternal weight change in the 2 years prior to pregnancy. Using this cohort, we will determine associations between preconception weight change rates on pregnancy, infant, and longer term maternal and child outcomes. Specifically, we will test the study’s central hypothesis that preconception weight change toward a normal BMI (weight loss in women with high BMI; weight gain in women with low BMI) reduces risk of adverse pregnancy and birth outcomes (GDM, gestational hypertension/preeclampsia, cesarean section, preterm birth, small for gestational age) by improving pre-pregnancy weight status. However, we also hypothesize that preconception weight loss may increase GWG, increasing risks of outcomes sensitive to GWG (postpartum weight retention, large for gestational age, infant weight gain, child BMI). We further hypothesize that these associations are strongest among women with more extreme BMI (underweight, Class II-III obesity).

The MatTrack cohort will advance our understanding of how preconception weight change impacts pregnancy, postpartum, and child outcomes in several ways. First, the MatTrack study is designed to inform weight management recommendations within a clinically relevant time period of approximately 2 years. In contrast, most prior studies examined associations between weight changes throughout a longer period prior to pregnancy, starting as early as 7 years of age, and subsequent health outcomes. [23–31] Second, we leverage clinically measured weights recorded longitudinally in the EHR, improving on prior evidence, which was predominately based on retrospectively self-reported weights. [10, 25, 27–30] Third, the MatTrack cohort will enable examination of associations with incremental rates of weight loss or weight gain. Prior observational studies examined variable weight loss definitions, such as self-reported categories of weight change (e.g., ≥5 kg loss, weight stable, ≥5 kg gain), [10, 23–25, 27, 28, 30, 32] or changes in BMI category from one time to the next (e.g., overweight in adolescence, normal weight in adulthood). [26, 31]

With regard to outcomes, the high retention and clinical engagement of patients throughout and after pregnancy, as well as linkage of clinical data from children, will enable the examination of longer-term maternal and child outcomes, for which evidence is scant. [29, 33] The extensive clinical data enables objective ascertainment of study outcomes and comorbidities, such as laboratory data to identify GDM and ICD-9 and ICD-10-CM codes to identify maternal comorbidities or child conditions. Lastly, our large study population enables examination of associations within starting BMI subgroups, including obesity class. In contrast, many prior studies examine all BMI categories combined [26, 28–31] or stratify by broad BMI categories that do not distinguish between obesity classes. [10, 23–25, 27, 32] We also have the ability to examine associations between preconception weight loss and outcomes among subgroups defined by both baseline BMI category and comorbidities.

Several limitations of the MatTrack cohort should be acknowledged. First, patients who were not insured during the time frame of interest were excluded, limiting generalizability to insured populations of U.S. patients with access to health care. Second, sufficient numbers of recorded weights were required in order to calculate preconception weight change, which may have enriched the cohort with patients with higher healthcare utilization due to greater clinical needs; however, starting BMI and clinical and sociodemographic characteristics were similar in those with repeated weight measures compared to the more inclusive cohort, reducing potential for selection bias. Third, while we are able to examine patient characteristics associated with various rates of weight change within the cohort, we are not able to assess patient intentionality related to weight change. Fourth, while we used BMI as a means to stratify patients for analysis, aligning with recommendations for women with elevated BMI to achieve a healthy weight prior to pregnancy, we recognize that BMI has poor sensitivity for identifying patients with excess adiposity, especially for people in intermediate ranges of BMI, and is inconsistently associated with health risks.[34, 35] Finally, while the cohort includes patients from four U.S. regions, findings may not be generalizable to the U.S. population.

With this well characterized cohort, we will report upon the association of preconception trajectories and the following outcomes: gestational weight gain, 6-week and 1-year postpartum weight retention, GDM, infant size for gestational age at birth, early childhood growth, and child BMI at 3 years of age. MatTrack study findings will examine the effects of objectively measured rates of preconception weight loss or gain on maternal and child outcomes extending beyond delivery, within important clinical subgroups. Findings will inform clinical understanding of the benefits and risks of weight change prior to pregnancy, allowing for more evidence-based preconception and interpregnancy care recommendations. Findings will also lay a foundation for future research examining other outcomes such as hypertensive disorders of pregnancy, early pregnancy loss, and longer-term outcomes such as metabolic conditions later in childhood or in the maternal life course.

## Data Availability

The datasets generated and analyzed during this study are not publicly available due to sourcing patient-level data from multiple health and data systems, which have restrictions regarding the availability and rerelease of data under cross-institution agreements. Data are available on reasonable request and with permission of all relevant parties from the KP Center for Health Research (contact investigator: Dr. Kimberly K. Vesco).

## Acknowledgements

The authors thank our study team of project managers: Daniel Sapp, Kaiser Permanente Center for Health Research; Mackenzie Kulik, KP Georgia, Center for Research and Evaluation; Johnathan Lai, Kaiser Permanente Hawaii, Center for Integrated Health Care Research; and Galina Inzhakova, Kaiser Permanente Southern California, Department of Research & Evaluation.

## Supporting information

**S1 File. growthcleanr Customizations**

**S1 Table. Baseline characteristics of pregnancies meeting different sets of MatTrack criteria**

**S2 Table**. **Preconception weight change rates (absolute weight change) by starting BMI category**

**S3 Table**. **Preconception weight change rates (as a percentage of starting weight) by starting BMI category**

